# Real-world impact of Elexacaftor-Tezacaftor-Ivacaftor treatment in young people with Cystic Fibrosis: A longitudinal study

**DOI:** 10.1101/2024.03.15.24304343

**Authors:** GJ Connett, S Maguire, TC Larcombe, N Scanlan, SS Shinde, T Muthukumarana, A Bevan, RH Keogh, JP Legg

## Abstract

**Introduction:** Elexacaftor, Tezacaftor, Ivacaftor (ETI) became available in the UK in August 2020 to treat people with Cystic Fibrosis (CF) aged > 12 years. We report a real-world study of clinical outcomes in young people treated with ETI at our CF centre within the first two years of its availability.

**Methods:** Participants aged 12-17 were identified within our clinic, with demographic data supplemented by the UK CF registry. Comprehensive outcome data spanning two years pre- and two years post-initiation of CFTR modulators were compiled from various local sources, including patient records, medication delivery logs, and clinical notes.

**Results:** Of the 62 patients started on ETI (32 male, mean age 13.3 years), most (76%) were homozygous for the F508del mutation. Three discontinuations occurred: one pregnancy, two related to side effects. Adherence was high (Proportion of Days covered >90% both years). Following ETI initiation there was a significant increase in mean FEV1% (+11.7 units; 95% CI 7.4 – 15.6), sustained throughout the two-year treatment period. There was no association between baseline lung function and the degree of improvement or rate of decline post-treatment. Improvements were similar for all treatable genotypes. There was a small increase in BMI z-score at four months of treatment, returning to baseline by 24 months. There was a marked reduction in the need for intravenous antibiotics.

**Conclusions:** ETI use in adolescents in a real-world setting led to sustained improvements in health outcomes, consistent with those seen in open trial extension studies

- **What is already known on this topic -** Clinical trials have demonstrated the efficacy of the highly effective CFTR modulator ETI in improving health outcomes for CF patients. However, there is a significant gap in understanding its real-world impact, particularly in young patients where adherence to optimise long-term outcomes is crucial.
- **What this study adds -** ETI provides sustained real-world benefits in young people with CF, including better lung function and reduced need for intravenous antibiotic treatment. High adherence likely plays a role.
- **How this study might affect research, practice or policy -** These findings support the widespread adoption of ETI in eligible CF patients and emphasise the need for further research to assess its long-term benefits and optimal integration into CF treatment protocols.

## Introduction

Cystic Fibrosis (CF) is one of the most prevalent life limiting autosomal recessive diseases. It primarily affects the respiratory and gastrointestinal systems. It is caused by Cystic Fibrosis Transmembrane Conductance Regulator (CFTR) gene variants that code for aberrant or absent CFTR protein [1]. The resultant impairment in epithelial chloride transport causes the accumulation of viscous airway mucus, chronic bacterial infection and inflammation [2].

Progressive bronchiectasis leads to respiratory failure and premature death[3]. Until recently, treatment has focused on the downstream consequences of abnormal CFTR function, but the advent of highly effective CFTR modulators has resulted in a transformative shift, targeting the underlying protein dysfunction. Elexacaftor, Tezacaftor, Ivacaftor (ETI), the most recent of these treatments, was first available in the UK in August 2020 for people with CF aged over 12 years with at least one copy of the DF508 gene variant or one copy from a list of other gene variants for which there was in vitro evidence of efficacy. Approximately 95% of the UK CF population have such a genotype and 61% of the UK CF population were receiving ETI treatment in December 2022 [4]. Clinical studies have confirmed ETI’s efficacy in improving lung function, quality of life, and reducing pulmonary exacerbations [5,6]. These benefits are also evident in those with advanced pulmonary disease [7].

Data for the real-world effectiveness of these treatments is limited. A longitudinal study by Mitchell et al. focusing on Ivacaftor in patients with the Gly551Asp gating mutation highlighted that whilst there were initial improvements in lung function, these were not sustained over a 5-year period [8]. Importantly, a diminished medicine possession ratio, indicative of reduced adherence, was associated with worse outcomes.

In this study we have investigated the real-world efficacy of ETI over the first two years of its availability in all young people aged 12-17 years and attending a single UK CF centre.

## Methods

### Patients

Young people aged under 18 years with CF and newly eligible for ETI in the first year of its availability after August 2020 were included. All were attending a single paediatric centre in the Wessex region of England with demographic data supplemented by the UK CF registry. All were aged over 12 years before starting ETI except one aged 11 years who started treatment on compassionate grounds. Three additional patients included in this study had started ETI prior to August 2020 having participated in the open trial extension study of this new treatment. All patients who met eligibility criteria were included in the analysis on an intention to treat basis.

### Data collection

Clinical data were extracted from patient records supplemented with data from the UK CF registry spanning a 2-year period before having ever received a CFTR modulator and for 2 years after starting ETI. This was not necessarily a continuous 4-year period. Eligible patients with two copies of the DF508 gene variants had been on Tezacaftor/Ivacaftor for up to 7 months before the end of their pre-ETI and ETI periods.

Baseline demographics recorded at the start of the 2-year pre-ETI period included age, sex, FEV1% (GLI reference data), Body Mass Index (BMI) z-score (WHO reference data) and CFTR genotype.

FEV1% measurements and growth data were obtained at the time of routine face to face clinic visits. These occurred once every 4 months during both the pre-ETI and ETI periods according to standard practice for all our CF patients. Lung function and growth data were obtained more frequently if patients made additional clinic visits because of concern about increased respiratory symptoms.

Other clinical data recorded at clinical visits included use of regular inhaled steroids, nebulised DNase, nebulised hypertonic saline, and proton pump inhibitors, pancreatic status (based on faecal elastase levels), use of enteral feeding, dose of oral lipase supplements, and HbA1c levels.

### Adherence (PDC-Proportion of Days Covered)

Patients accessed ETI through a homecare delivery company. The company used a monthly contact protocol to monitor each person’s medicine stock. This was to help ensure that they could provide uninterrupted ETI when further supplies were requested.

Adherence was estimated using Proportion of Days Covered (PDC), a validated metric derived from pharmacy records [9]. A PDC was calculated for the first and second year of ETI treatment.

### Statistical analysis

The highest FEV1% and BMI z-score within each four-month period were used in the analysis to mitigate the effect of short-term fluctuations in health. Each patient was therefore assigned a single measurement for each 4-month period.

PDCs for years 1 and 2 of ETI were summarised by sex and genotype (heterozygous or homozygous for ETI responsive gene variants). Four monthly mean FEV1% and mean BMI z-score were plotted over time. We estimated the association of ETI treatment with the step-change in FEV1% over the first 4 months of treatment, and with the slope of FEV1% over the subsequent 2-year follow-up period. To take into account the repeated measures of FEV1%, using data from both the pre-ETI period and the treatment period, the analysis used a generalised estimating equations (GEE) approach with an independent working correlation matrix. The model relates the expected FEV1% at time *t* months after baseline (*t* = 4,8,12,16,20,24) to ETI status, *t*, and their interaction, plus covariates sex, baseline FEV1% and baseline age. In the pre-ETI period ‘baseline’ (*t* = 0) refers to the start of the 2-year pre-ETI period, and in the treatment period ‘baseline’ (*t* = 0) refers to the start of the 2-year treatment period. The same analysis was performed for the BMI z-score outcome, replacing the adjustment for baseline FEV1% with adjustment for baseline BMI z-score. These models were used to estimate the typical trajectory of FEV1% and BMI z-score over time in the two time periods. To quantify the effect of PDC on FEV1%, PDC was added to the model and fit on the ETI period only. The PDC model coefficient is the estimated association between PDC and FEV1%.

All other clinical outcomes were compared in the pre-ETI and ETI periods. There were two measurements for binary outcomes – recorded at the end of the pre-ETI period, and at the end of the ETI period. These included regular inhaled steroid use, regular mucoactive use, proton pump inhibitor use, pancreatic status, use of enteral feeds, insulin dependent diabetes, liver disease (receiving treatment with ursodeoxycholic acid or CF related liver USS changes) azithromycin prophylaxis, and use of regular nebulised antibiotics. For ‘count’ outcomes (total days of IV antibiotics, number of ABPA episodes, number of *Pseudomonas aeruginosa* isolates, number of *Staphylococcus aureus* isolates) we compared the rates per person per year. We also considered the number of courses of oral antibiotics per patient per year as a categorical variable. Finally, we measured pancreatic enzyme dose (lipase/kg/day) and HbA1c in the second year of the pre-ETI and ETI periods as continuous variables. Tests of null hypothesis of no differences between these outcomes in the two periods were performed. P-values for tests of continuous, binary, count, and categorical outcomes were obtained based on paired t-tests, McNemar’s test, Poisson regression with robust standard errors, and Friedman’s test respectively. Statistical analyses were performed using R V4.2.1 [10]. GEEs were fitted using the geepack package [11].

## Results

### Patient Demographics

Sixty-two young people were eligible for and commenced treatment with ETI. The mean age at starting treatment was 13.9 years (range 11 years – 16.6 years). Baseline demographic and clinical characteristics for the pre-ETI and ETI periods are summarised in Table 1. Fifty-nine young people remained on treatment after two years. Treatment with ETI was terminated in one patient due to pregnancy, and in two patients because of adverse effects. The number of patients providing data at each 4 monthly clinic visit is shown in Table 2.

**Table 1:** Summary of baseline participant characteristics at the start of the pre-ETI and ETI periods.

| Baseline measurements | Start of Pre-ETI period | Start of ETI period |
| --- | --- | --- |
| Age (years) [Mean (range)] | 10.9 (8.6-15) | 13.9 (11-16.6) |
| BMI (z-score) [Mean (SD)] | +0.05 (0.86) | +0.1 (1.01) |
| FEV1% [Mean (SD)] | 86.54 (19.68) | 82.42 (18.87) |
| *Patient characteristics* | N (%) |  |
| Sex – Male | 32 (51.6%) |  |
| Genotype - 2 ETI responsive variants | 47 (75.8%) |  |
| Genotype – 1 ETI responsive variant | 15 (24.2%) |  |

**Table 2:** Number of observations of FEV1% and BMI z-score over time.

|  | Pre-ETI modulator Period |  |  |  |  |  |  | ETI Period |  |  |  |  |  |
| --- | --- | --- | --- | --- | --- | --- | --- | --- | --- | --- | --- | --- | --- |
| Time (months) | -24 | -20 | -16 | -12 | -8 | -4 | 0 | 4 | 8 | 12 | 16 | 20 | 24 |
| FEV1% | 59 | 60 | 60 | 59 | 58 | 60 | 52 | 61 | 53 | 58 | 55 | 52 | 51 |
| BMI z-score | 61 | 61 | 61 | 61 | 61 | 62 | 62 | 62 | 62 | 62 | 61 | 58 | 47 |

Month 0 is baseline measurements immediately before starting ETI treatment. Positive months are on treatment and negative months are in the observation period. This is not necessarily a continuous 4-year period, as some patients in the study were on Tezacaftor/Ivacaftor for up to 7 months between the end of their observation period and starting ETI treatment.

### Adherence

Adherence to the ETI regimen remained robust across the study duration (Table 3). Adherence was higher in year 1, with a marginal, non-significant reduction in year 2. Even after this slight reduction, the mean ETI adherence remained above 90% in year 2. A 1% increase in ETI adherence in the ETI period was associated with an increase of 0.25 (SE = 0.16) FEV1%, but this association was not significant (p = 0.11).

**Table 3:** Mean ETI adherence in years 1 and 2 after initiating ETI, defined as the percentage of days covered. P-values calculated using paired t-test.

|  | **Year 1**<br>**Mean (SE)** | **Year 2**<br>**Mean (SE)** | **Mean**<br>**difference** | **P-value** |
| --- | --- | --- | --- | --- |
| **Overall** | **94.4 (9.9)** | **90.7 (17.0)** | **-3.5** | **0.07** |
| **Males** | **96.1 (7.8)** | **92.9 (15.8)** | **-3.2** | **0.20** |
| **Females** | **92.6 (11.9)** | **90.9 (12.3)** | **-1.3** | **0.50** |
| **Genotype - 2 ETI**<br>**responsive variants** | **94.4 (9.8)** | **90.8 (17.9)** | **-3.2** | **0.10** |
| **Genotype – 1 ETI**<br>**responsive variant** | **94.4 (10.6)** | **90.3 (14.2)** | **-4.1** | **0.30** |

### Lung function

There was a significant increase in FEV1% (11.7 units; 95% CI 7.4 – 15.6) from baseline in month 4 after starting ETI treatment. At the end of the study improvements in FEV1% were well sustained. The rate of FEV1% decline was slower after the initiation of treatment, although this difference was not significant (p-value for interaction term = 0.48). Figure 1 shows changes in mean FEV1%. Table 4 shows the estimated slopes for FEV1% trend lines, adjusted for age and genotype in the pre-ETI period and from 4 months after starting ETI.

**Table 4:**
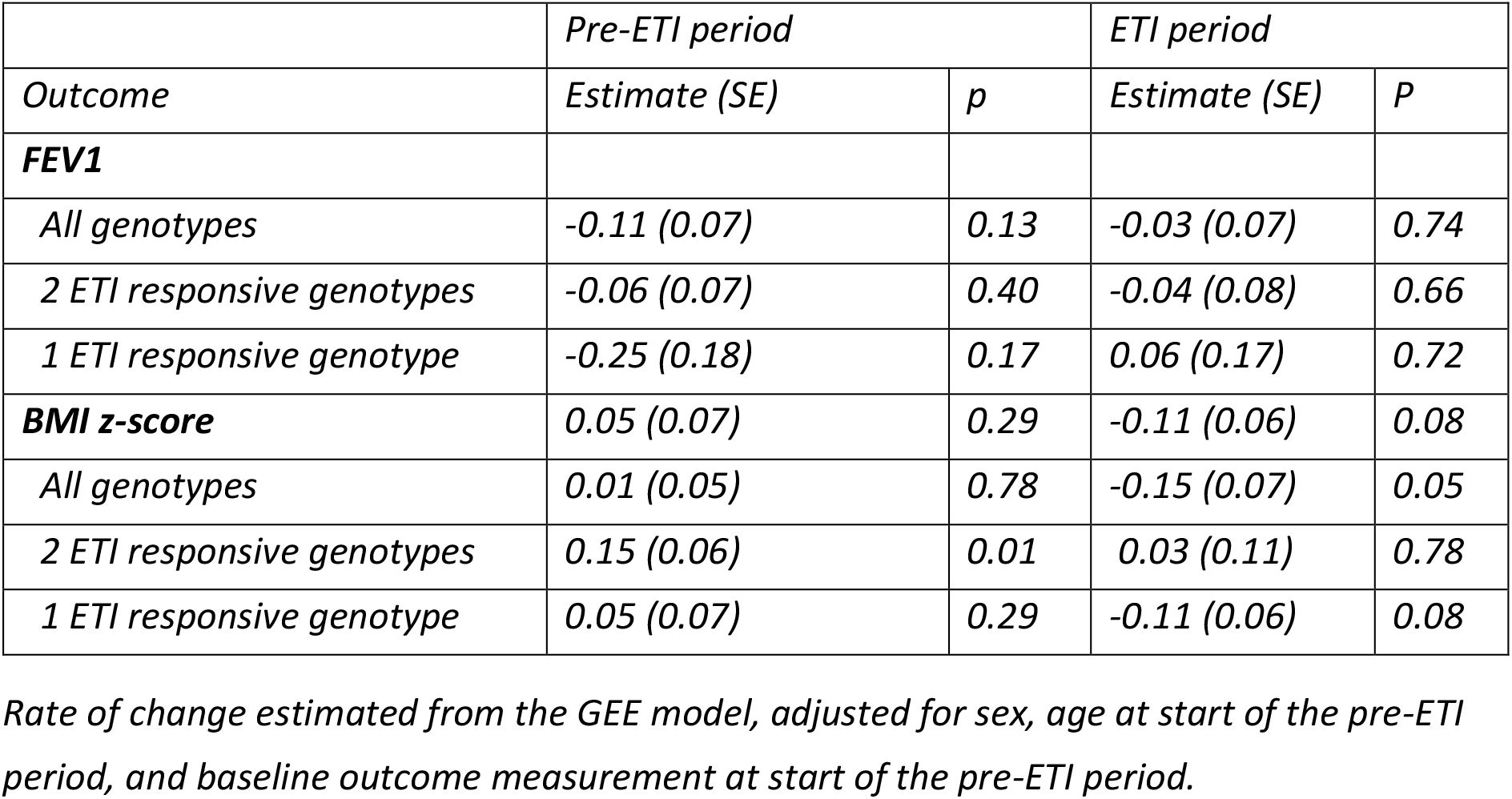
Association between ETI treatment and rate of FEV1% and BMI z-score change by genotype.

**Table 4:** Association between ETI treatment and rate of change in BMI z-score over time.

|  | pre-ETI period |  | ETI period |  |
| --- | --- | --- | --- | --- |
| Genotype | Estimate (SE) | p | Estimate (SE) | p |
| All genotypes | 0.05 (0.07) | 0.29 | -0.11 (0.06) | 0.08 |
| 2 ETI responsive variants | 0.01 (0.05) | 0.78 | -0.15 (0.07) | 0.05 |
| 1 ETI responsive variant | 0.15 (0.06) | 0.01 | 0.03 (0.11) | 0.78 |

**Figure 1:**
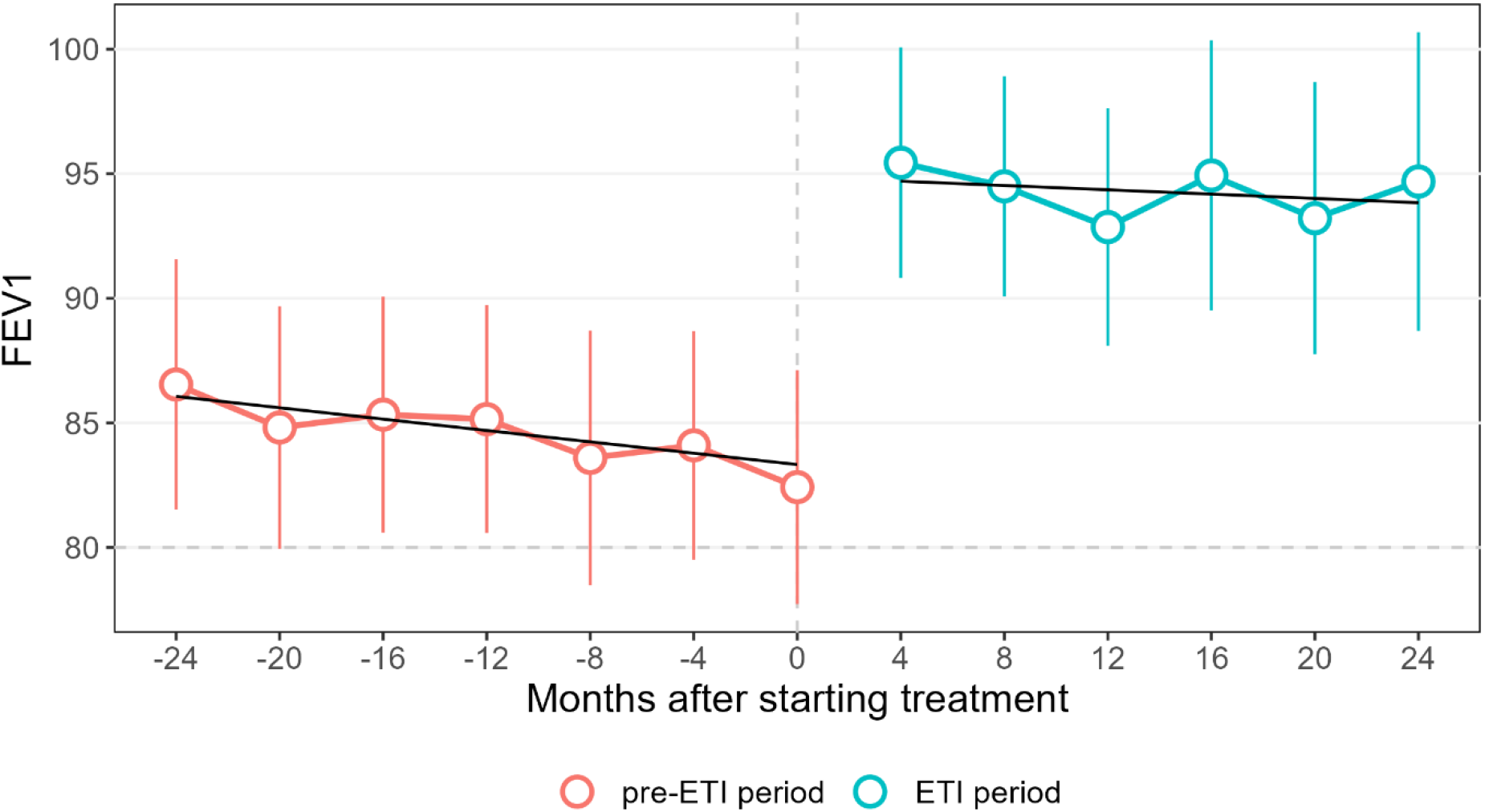
Changes in Mean FEV1% Pre- and Post ETI treatment. *Error bars are 95% confidence interval for the means. Black lines are marginal mean predicted values from a GEE model adjusting for age, sex, and FEV1% at the start of each period*.

### Body Mass Index

Body Mass Index z-score increased significantly when measured 4 months after starting ETI treatment (0.25 units; p = < 0.001). This small increase was not sustained and by the end of two year’s treatment mean BMI z-score was similar to baseline values (Figure 2).

**Figure 2:**
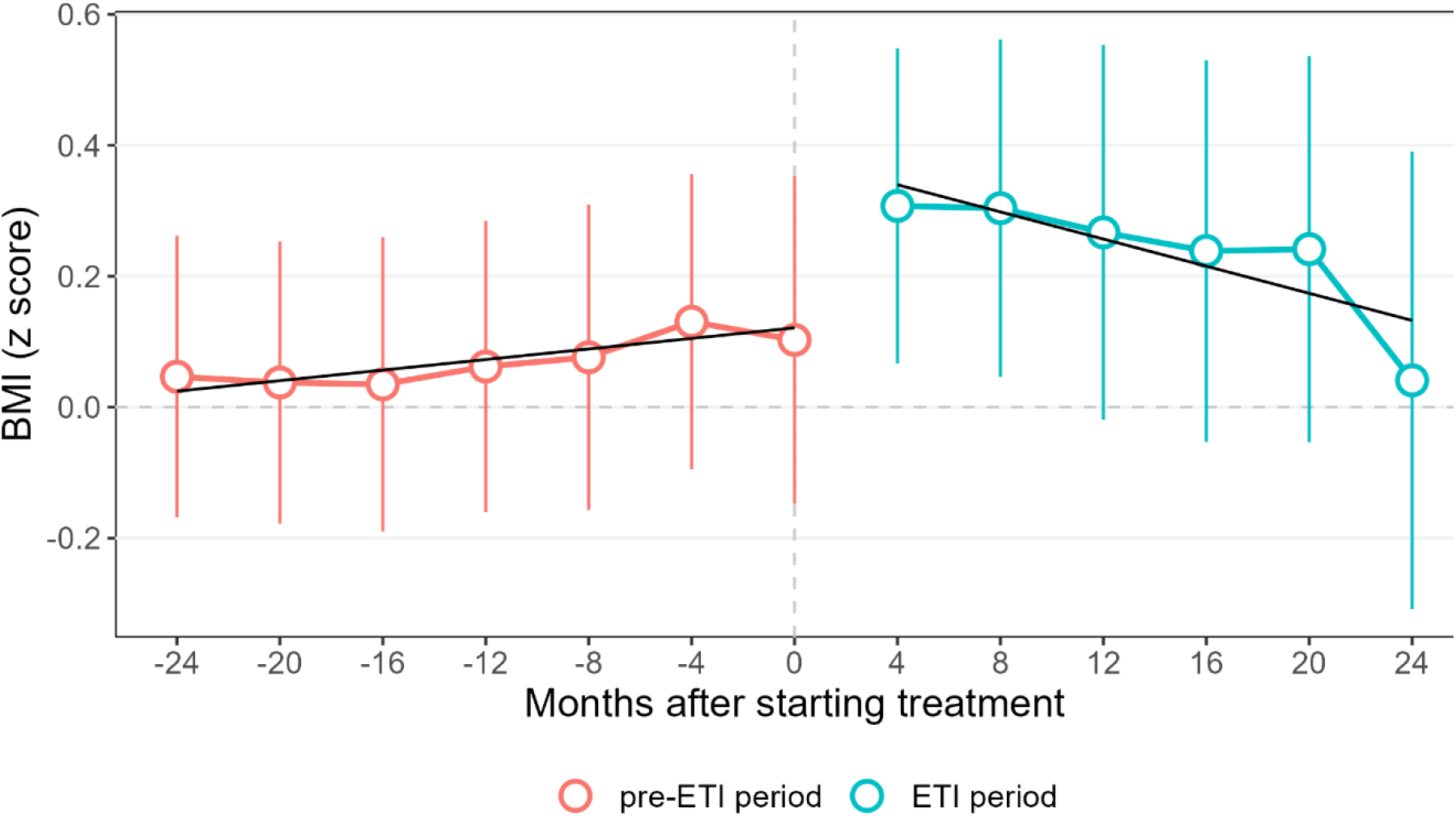
Changes in Mean BMI Z-score Pre- and Post ETI treatment. *Error bars are 95% confidence intervals. Black lines are the marginal mean predicted values from a GEE model, adjusting for age, sex, and BMI z-score measured at the start of each period. The dotted horizontal line indicates the 50*^*th*^ *Centile for BMI z-score as per World Health Organization standards*.

As shown in table 4, there were no significant changes in nutritional status as a result of ETI treatment either at a population level or by genotype.

As shown in table 4, there were no significant changes in nutritional status as a result of ETI treatment either at a population level or by genotype.

### Clinical Outcomes

A significant reduction in antibiotic usage occurred post ETI (Table 5). Most notable was an 88% reduction in the median number of days/year of IV antibiotics. There was also a significant decline in the number of isolates of the key pathogens *Pseudomonas aeruginosa* and *Staphylococcus aureus*, suggesting that ETI was significantly impacting on the bacterial load within the airways.

**Table 5:** Comparison of clinical outcomes for pre-ETI and ETI treatment periods.

|  | N | Pre-ETI period | ETI period | P-value |
| --- | --- | --- | --- | --- |
| **Antibiotic use** |  |  |  |  |
| Azithromycin Prophylaxis, n (%) | 62 | 15 (24%) | 9 (15%) | 0.08 |
| Regular nebulised antibiotics, n (%) | 61 | 41 (67%) | 16 (26%) | <0.01 |
| Total number of days receiving IV antibiotics, median (range for number of days treated) | 62 | 32 (7-64) | 4 (3-28) | <0.01 |
| Number of courses of oral antibiotics / patient / year | 35 |  |  | <0.01 |
| None |  | 4 | 18 |  |
| 0.5 to 1 |  | 4 | 7 |  |
| 1.5 to 2 |  | 11 | 7 |  |
| 2.5 to 3 |  | 7 | 3 |  |
| >3 |  | 9 | 0 |  |
| Insulin dependent diabetes, n (%) | 62 | 13 (21%) | 15 (24%) | 0.62 |
| Liver disease, n (%) | 62 | 21 (34%) | 18 (29%) | 0.45 |
| **Microbiology** |  |  |  |  |
| ABPA, n | 62 | 9 | 1 | 0.01 |
| Pseudomonas aeruginosa, n | 62 | 34 | 10 | <0.01 |
| Staphylococcus aureus, n | 62 | 41 | 23 | <0.01 |
| **Other clinical outcomes** |  |  |  |  |
| Regular inhaled steroids, n (%) | 62 | 26 (42%) | 8 (13%) | <0.01 |
| Regular mucoactives, n (%) | 62 | 60 (97%) | 52 (84%) | 0.01 |
| Proton pump inhibitors, n (%) | 62 | 34 (55%) | 22 (36%) | <0.01 |
| Pancreatic sufficiency, n (%) | 62 | 2 (3%) | 2 (3%) |  |
| Enteral feeding, n (%) | 62 | 7 (11%) | 2 (3%) | 0.07 |
| Lipase (units/kg/day), mean (SD) | 56 | 15,900 (12,100) | 9,500 (6,200) | <0.01 |
| HbA1c, mean (SD) | 33 | 41.4 (12.3) | 40.2 (15.2) | 0.15 |

*Liver disease refers to patients receiving treatment with Ursodeoxycholic acid either for CF related liver USS changes and/or persistently raised liver function enzymes. P-values for tests of continuous, binary, count, and categorical outcomes calculated using paired t-tests, McNemars test, Poisson regression with robust standard errors, and Friedmans test respectively.*

Amongst the continuous outcomes, there was a significant reduction in median lipase dose. This was probably a result of improved intestinal function because pancreatic lipase levels when measured were consistently below the lower levels of detection irrespective of changes in the need for enzymes. There was a slight non-significant decrease in HbA1c levels which was reassuring given that glucose homeostasis tends to get worse over time in this age group.

## Discussion

The introduction of highly effective modulator therapy has revolutionised the management of CF. This real-world study longitudinally assessed the clinical efficacy of ETI in a cohort of young people with CF, focusing on adherence and the impact on respiratory and gastrointestinal morbidity.

After initiating ETI, there was a significant increase in FEV1%, which remained consistent until the end of the observation period. These findings are in keeping with the sustained improvements in lung function reported in the open trial extensions of phase 3 clinic trials[12]. There were no obvious differences in clinical outcomes between individuals with one or two copies of gene variants for which there was evidence of ETI responsiveness.

Higher levels of adherence to ETI were associated with a slower decline in FEV1%, although the association was not significant. Overall adherence as estimated by proportion of days covered was high. Whilst this was consistent with sustained improvements in lung function and decreased burden of care, it meant that we could not determine whether poor adherence was associated with less good outcomes. Our findings are consistent with data from the phase 3 clinical trials of ETI [5,13,14] and the open-label extension trial data conducted over a similar time frame[12]. Our short-term observations are also consistent with other real-world studies of similar age groups reporting outcomes over a 12 month period[15–17].

Our findings are in contrast to the real world study of ivacaftor reported by Mitchell *et al*. in which they saw a significant decline in lung function that was obvious within the first 2 years of treatment[8]. Our study included a younger entirely paediatric population when treatment was first started (mean age 13.9 years versus 29.1 years) with better mean lung function prior to starting modulator treatment (FEV1% 82.4 versus 58.4). These differences in age and disease severity as well as better adherence are likely to be important factors influencing treatment response[18–20]. Our results are likely to be subject to the influence of parental supervision on treatment adherence [21,22]. It is also worth noting that ETI is a more effective modulator as demonstrated by its superiority over ivacaftor in treating gating mutations [15].

CFTR modulators have been shown to be effective in enhancing weight and BMI. Our data showed a small but significant increase in BMI z-score of 0.25 units four months after starting ETI, but this was not sustained. This short term effect is consistent with the findings of Heijerman et al, who observed a 0.6 kg/m^2^ increase in BMI four weeks after initiating ETI treatment in Phe508del homozygous patients [14]. Mainz et al. in a cohort of 107 children demonstrated a more sustained increase in BMI z-score of 0.42 units, up to 22 weeks after starting ETI treatment[23]. In our study the mean BMI z-score 24 months after starting ETI therapy was very similar to that at baseline.

There are several possible explanations for this. A previous study examining changes in BMI after starting ETI treatment highlighted that the most substantial weight gain tends to occur in individuals with lower body weights [24]. Our CF population had a healthy mean BMI z-score at the start of the study of +0.1. The mean age when starting ETI therapy was 13.9 years and many individuals had yet to experience their pubertal growth spurt. Should this have occurred within the two-year period following initiation of ETI therapy, it is possible that any spike in BMI-z score would be offset by a significant increase in height which is typically accompanied by a plateau in weight gain.

Our relatively frequent clinical reviews are also likely to have had an effect. It has been reported that increased compliance with weight management strategies and the adoption of healthier lifestyles are more likely to be achieved through more frequent follow-up interventions [25]. Whilst intervening early with dietary advice and dispelling historical beliefs around high energy diets in CF was key, our comprehensive approach to weight management is further enriched by collaborative work with our dieticians, physiotherapists, exercise physiologists and social worker to achieve a more holistic strategy for nutritional outcomes[26]. Annual blood tests indicated that most patients did not exhibit malabsorption of fat-soluble vitamins and in many cases doses of Vitamin A and E could be reduced.

Our observed decrease in antibiotic use is consistent with results from previous studies reporting similar trends after starting highly effective modulator treatment [13,27]. This will likely mitigate against the risks of long term antibiotic resistance across the life course [28]. The significant reduction in the use of inhaled steroids and mucoactive nebulisers is consistent with evidence that ETI reduces airway inflammation and improves mucociliary clearance [29,30].

The decline in the number of isolates of *Pseudomonas aeruginosa* and *Staphylococcus aureus* is consistent with evidence that ETI treatment can reduce the bacterial pathogen load within the airways [31,32]. However, many patients were less able to expectorate sputum after starting ETI and this would have impacted on the quality of specimens obtained for microbial culture.

Our study has several limitations. It was a single centre study which might limit the generalisability of our findings to a broader CF population. The relatively short follow-up period of 2 years might not be sufficient to capture the full extent of long-term outcomes after starting ETI and longer-term studies are clearly needed. Our data collection period partially coincided with the COVID-19 pandemic. Although our clinical surveillance routine remained consistent throughout this period with 4 monthly face to face patient contact, the effects of lock downs and changes in patient behaviour would likely have resulted in fewer episodes of infection and better adherence to daily treatment regimes.

## Conclusion

This single centre study has demonstrated significant clinical benefits after ETI treatment in a real-world setting. Sustained respiratory improvements and a positive effect on other clinical outcomes were readily apparent. Our findings demonstrate how ETI can positively impact on the disease trajectory in young people whilst transitioning to adult services.

Future studies might usefully validate these findings in larger multi-centre studies over a longer period. Strategies to maintain high levels of adherence in young people will be critical to realising the full potential of modulator treatment.

## Data Availability

All data produced in the present work are contained in the manuscript

## Acknowledgements

We thank people with cystic fibrosis and their families for consenting to their data being held in the UK CF Registry. The UK CF Registry is sponsored and coordinated by the UK Cystic Fibrosis Trust.

## Contributors

GJC and JPL conceived the study format. GJC, JPL, TCL, NS, SSS, TL and AB contributed to the data acquisition and set-up. RHK and SM conceived and conducted the statistical analysis.

JPL and GJC drafted the manuscript with substantial input on presentation and interpretation of statistical methods and results RHK and SM. All authors approved the final version of the manuscript and agree to be accountable for all aspects of the work.

## Patient consent for publication

Not required.

## Ethics approval

This work used anonymised data from the UK Cystic Fibrosis Registry, which has Research Ethics Approval (REC ref: 07/Q0104/2). The use of the data was approved by the Registry Research Committee (Data Request Reference 382). Data are available following application to the Registry Research Committee. https://www.cysticfibrosis.org.uk/the-work-we-do/uk-cf-registry/apply-for-data-fromthe-uk-cf-registry. This work was also approved by the London School of Hygiene & Tropical Medicine Research Ethics Committee (Ref 21866).

